# Peripherin: a novel early diagnostic and prognostic plasmatic biomarker in Amyotrophic Lateral Sclerosis

**DOI:** 10.1101/2025.01.24.25321038

**Authors:** Alessandro Bombaci, Giovanni De Marco, Federico Casale, Paolina Salamone, Giulia Marchese, Giuseppe Fuda, Andrea Calvo, Adriano Chiò

**Affiliations:** IRCSS Policlinico San Donato, San Donato Milanese, Italy; Vita-Salute San Raffaele University, Milan, Italy; Turin ALS Centre, “Rita Levi Montalcini” Department of Neuroscience, University of Torino, Turin, Italy; SC Neurologia 1U, AOU Città della Salute e della Scienza di Torino, Turin, Italy

**Keywords:** fluid-biomarker, HSP, PLS, PRPH, neurofilament

## Abstract

**Background and aim:** Motor neuron diseases (MND) are heterogeneous and complex neurodegenerative disorders. Biomarkers could facilitate early diagnosis, prognosis determination, and patient stratification. Among the most studied biomarkers are neurofilaments, with peripherin (PRPH), a specific type predominantly expressed in peripheral nervous system neurons, gaining attention. To date, no studies have evaluated PRPH levels in human plasma.

**Methods:** sandwich-ELISA was used to quantify plasma peripherin from 120 MND (100 ALS, 4 PMA, 15 PLS), 73 MND-mimics (including myelopathy, radiculopathy, axonal neuropathies, Hirayama disease, IBM, benign fasciculation syndrome, functional syndrome, myasthenia, post-polio syndrome) and 38 healthy-controls (HCs). Plasma was collected at the time of diagnosis or some months earlier. 41 ALS were evaluated longitudinally. ALSFRSr, MRC, spirometry, genetic tests, disease progression rate (PR), blood examinations, and neuropsychological tests were performed. Statistical analyses included Kruskal-Wallis, Mann-Whitney, Cox-regression, and Kaplan-Meier curves.

**Results:** PRPH plasma levels is different between groups (p<0.0001); Bonferroni’s correction shows higher PRPH in MND compared to MND-mimics and HCs. Comparing PLS with HSP PRPH resulted to be higher (p=0.0001). Differences are confirmed co-variating for age and sex. ROC curve shows a good capability of PRPH to discriminate ALS from MND mimics (AUC= 0.85).

PRPH levels correlated directly with ALSFRSr, and with lower body involvement while inversely with PR. Higher PRPH levels were associated with survival advantages.

**Discussion:** Plasma PRPH levels are elevated in MND, particularly ALS, from the early disease stages, distinguishing MND from mimics and correlating with clinical parameters and survival. Further multicentre studies incorporating additional biomarkers are required to refine the diagnostic and prognostic utility of PRPH in MND.

## Introduction

Motor neuron diseases (MNDs) encompass a group of fatal neurodegenerative disorders marked by progressive motor neuron degeneration^1^. Amyotrophic lateral sclerosis (ALS) is the most common form, involving both upper and lower motor neurons and leading to weakness in bulbar, limb, trunk, and respiratory muscles^2^. Approximately 10% of ALS cases are familial (fALS), while the remaining 90% are sporadic (sALS)^3^. Cognitive or behavioural impairments affect up to 50% of ALS patients, and approximately 10% exhibit frontotemporal dementia (FTD).^1^ Despite extensive research, the aetiology and pathophysiology of ALS remain unclear, with a median survival of about three years post-diagnosis. Other MNDs include primary lateral sclerosis (PLS), involving upper motor neurons, and progressive muscular atrophy (PMA), which primarily affects lower motor neurons.

Identifying diagnostic and prognostic biomarkers is crucial for differentiating ALS from similar conditions, stratifying patients, predicting disease progression, and improving clinical trial inclusion. Neurofilaments (NFs) have emerged as promising biomarkers for ALS and other neurodegenerative diseases^4,5^. Beyond the widely studied neurofilament triplet proteins (NFTPs, i.e. Neurofilament Light Chain [NfL], Neurofilament Medium Chain [NfM], and phosphorylated- Neurofilament Heavy Chain [pNfH]), and alpha-internexin, another type III intermediate filament belong to the group of neurofilaments, the peripherin (PRPH). It forms heteromers as a fourth subunit in the peripheral nervous system’s neurofilaments^6^. As well as all the other NFTPs, PRPH present a stoichiometry, fundamental for an adequate axonal transport and for an appropriate cell structure^7^. Alterations to this stoichiometric balance of intermediate filament is associated with the formation of cytoplasmatic inclusions in motor neurons of ALS-models^8^. This phenomenon seems to be the result of the loss of proper regulation of intermediate filament mRNA metabolism^7,9^. Moreover, PRPH is fundamental not only in axonal transport and constitution of neuron cytoskeleton, but it fulfils a role in axonal development and in neuronal cells’ differentiation^10^.

Recent studies have explored PRPH in human biofluids, highlighting its potential as a diagnostic marker. Elevated PRPH levels in serum and cerebrospinal fluid (CSF) have shown promise in distinguishing ALS from dementia, spinal bulbar muscular atrophy (SBMA), and polyneuropathies.^11^ Another study demonstrated its utility in early Guillain-Barré syndrome diagnosis.^12^

Given the pivotal role of PRPH in motor neuron health and response to stress, this study investigates PRPH levels in the plasma of MND patients. We aim to elucidate its diagnostic and prognostic value, potentially enhancing our ability to differentiate ALS from other neurodegenerative and neuromuscular conditions.

## Materials and Methods

### Design of the study and clinical assessment

Firstly, we set up a preliminary test to validate the Human Peripherin (PRPH) ELISA Kit (Abbexa, Cambridge, UK) for measuring PRPH in human biofluids and determining the optimal dilution factor for ALS patients.

Secondly, we realised a retrospective, longitudinal study on a cohort of 192 patients and 38 healthy controls (HCs) referring to the Department of Neurosciences of the University of Turin (Italy), from June 2019 to March 2023, who underwent blood collection for diagnostic purposes. Blood samples were collected from MND patients at diagnosis and, for 41 ALS patients, again after six months. Participants with active cancer, infections, or autoimmune disorders were excluded.

Demographic and clinical data are summarised in **Table 3**, ensuring robust patient characterisation and comparability.

ALS patients received their diagnosis following the Gold Coast criteria^13^. Disease severity was assessed using ALS Functional Rating Scale-revised (ALSFRSr)^14^. Subscores included: ALSFRSr_noresp (excluding respiratory items), ALSFRSr_B (bulbar score: items 1 and 3), and ALSFRSr_4limb (limb score: items 4–9).

Moreover, all ALS patients underwent: a clinical evaluation (neurological examination, including the Medical Research Council scale [MRC]^15^ measurement, and ALSFRSr scale) at the moment of diagnosis, at the moment of blood sampling (T0), and after 6 months from sampling (T6), an instrumental evaluation (electromyography, cerebral MRI with tractography and functional-MRI, spirometry), a neuropsychological evaluation^16^, and a genetic test for the most common mutation (*C9orf72, SOD1, TARDBP* and *FUS* genes).

Patients were categorised by phenotype (classic ALS, bulbar ALS, respiratory ALS, flail arm, flail leg, and predominant upper motor neuron ALS).^2^ Progression rate of disease (PR) and progression rate without respiratory items (PR_noresp) at T0 were calculated as follow:

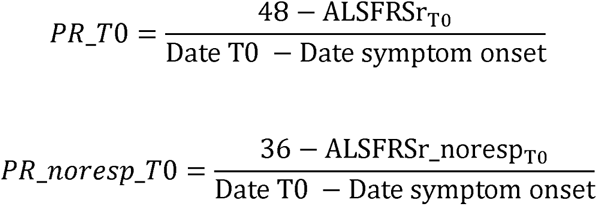

Disease progression rate and PR_noresp were calculated at T1 were calculated as follow:

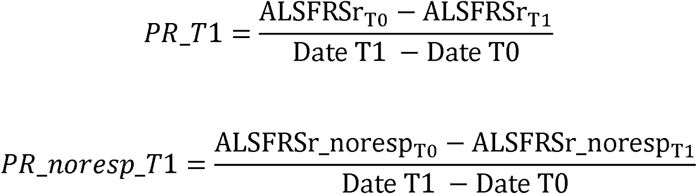

Moreover, we calculated in ALS patients an index of upper motor neuron (UMN) impairment, called UMN burden score (UMNBS), ranging from 0 to 24, collecting retrospectively data on burden and atrophy distribution from medical records of our Centre (**Table 1**). This is a readaptation of the Penn Upper Motor Neuron Score (PUMNS)^17^, based on data availability. We rated limb spasticity using the Modified Ashworth Scale (MAS)^18^.

**Table 1.** Upper Motor Neuron Burden Score (UMNBS)

|  |  |
| --- | --- |
| **Limb reflexes**<br>*(bicipital, brachioradialis, patellar, ankle)* | 0 if absent, reduced or normal in a normotrophic area<br>1 if brisk or retained in an atrophic area |
| **Pathological reflexes**<br>*Palmomental reflex (bilateral), Hoffmann's sign (bilateral), Babinski's sign (bilateral)*<br>Jaw jerk | 0 if absent<br>1 if present<br>0 if absent<br>1 if present<br>2 if brisk |
| **Spasticity at four limbs** (as PUMNS) | 0 if normal muscle tone<br>1 if MAS 2-3<br>2 if MAS 4-5 |

We also calculated a lower motor neuron index (LMNI; **Table 2**), implementing the scoring system proposed by Devine et al.^19^, whose score range between 0 to 12 points, and adding the bulbar lower motor neuron score^20^, whose score range between 0 to 3 points. The resulting LMNI consists in a complete scale able to evaluate the lower motor neuron involvement both in bulbar and in limbs regions, scoring between 0 to 15 points.

**Table 2.** Lower motor neuron index (LMNI)

|  |  | **Lower Motor Neuron Signs** |
| --- | --- | --- |
| **Limb Score (for each limb)**<br>[0-12] | 0 | No clinically significant involvement |
| | 1 | Definite, but trace involvement<br>Weakness $\geq 4/5$, involving one or more segments (with no segments $< 4/5$), and mild wasting |
| | 2 | Moderate involvement<br>Weakness $\geq 3/5$, involving one or more segments (with no segment $< 3/5$), and moderate wasting |
| | 3 | Significant and severe involvement<br>Little or no movement (LMN weakness $\leq 2/5$) involving |
|  |  | one or more segments, and severe wasting |
| Bulbar Score<br>[0-3] | 0 | No clinically significant involvement |
|  | 1 | Tongue atrophy and/or fasciculations |
|  | 2 | Score (1) + tongue hypomobility |
|  | 3 | Score (2) + jaw and/or face weakness |

Stage of disease was assessed using the two most known system: the KINGS^21^ and the MiToS^22^ staging systems.

### Laboratory evaluation

Plasma samples for preliminary kit evaluation were collected in EDTA tubes, centrifuged at 1500× g for 15 minutes at 18°C within one hour of collection, and stored at −80°C. Samples were anonymised using unique codes accessible only to authorised researchers. Before ELISA testing, samples were thawed at 4°C overnight, brought to room temperature for 30 minutes, and centrifuged at 3000× g for 15 minutes to remove platelets and cellular debris, following the protocol previously described^23^.

### Assay

We used the Human Peripherin (PRPH) ELISA Kit ELISA commercial Kit (Abbexa, Cambridge, UK), following the manufacturer’s instructions.

Each plate contained calibrators (0.312-20 ng/ml) in duplicate and two samples with known concentration of PRPH, provided in each kit. Manufacturer declared a Sample recovery range after spiking of 91–105% and a linearity range of 90–116% in dilutions up to 1:8 both in serum and in plasma.

After preliminary analysis, useful for validation of this kit in our samples and led following protocols adapted from Andreasson et al.^24^, all samples were distributed on the plate using a dilution of 1:2 in 0.01 mmol/l of phosphate-buffered saline (as per kit recommendations), and measured in duplicate. The inter-assay and intra-assay coefficients of variance were all below 10%. According to manufacturer’s instructions, analytical sensitivity was set <0.156 ng/ml.

Plates were read using a CLARIOstar Plus plate reader (BMG LABTECH, Ortenberg, Germany), and standard curves were fitted using 4-parameter logistic regression with MARS data analysis software. The median intra-assay and inter-assay coefficients of variation were below 15% for all the assays.

General blood tests were performed by our hospital laboratory.

### Statistical analysis

A Shapiro–Wilk test showed that the data were not normally distributed.

Kruskal–Wallis and Mann-Whitney tests were used to compare the groups, with Bonferroni post-hoc adjustments applied when significant differences emerged. The correlations between PRPH levels and clinical, laboratory and instrumental parameters were calculated by the Spearman rank correlation (r_s_). Multiple regression analyses were led.

Kaplan Meier curves were generated in patients who underwent death or tracheostomy, also stratifying for age, sex and site of onset. Multivariate Cox-regression analysis with a backward stepwise method were led.

Wilcoxon signed-rank test was assessed for longitudinal analysis.

Statistical significance was set at p<0.05, and all calculations were performed using SPSS Statistics V29 (Chicago, IL, USA). Ethical approval was granted by the Turin ALS Centre’s Ethics Committee (Comitato Etico Azienda Ospedaliero-Universitaria Città della Salute e della Scienza, Torino) (n° 0011613, 03/02/2020), and all participants provided written informed consent.

## Results

### Preliminary evaluations in biofluids

Using the Human PRPH ELISA Kit, we tested various dilutions (from undiluted to 1:8) of CSF and plasma from 3 MND patients and 3 HCs. PRPH was undetectable in CSF due to levels below the assay’s detection threshold. However, consistent with PRPH’s peripheral nervous system distribution^6^, it was measurable in plasma, with the optimal dilution determined to be 1:2 or undiluted.

### Evaluation of plasma PRPH in MNS patients, MND-mimics and healthy controls

We conducted a retrospective, longitudinal investigation involving 119 patients living with MND (100 with ALS, 15 with PLS, 4 with PMA), 73 cases of MND-mimics (including myelopathies, radiculopathies, axonal neuropathies, inclusion body myositis, post-poliomyelitis syndrome, myasthenia gravis with bulbar onset, primary progressive aphasia, Parsonage-Turner disease, Hirayama disease, syringomyelia, progressive supranuclear palsy, benign fasciculation syndrome, functional disorder, and hereditary spastic paraplegia [HSP]; see **Supplementary data, Table 4**), and 38 HCs.

Among ALS 60 were male (58%), 36 had a bulbar onset of disease (35%), and 2 a respiratory onset (2%). Among ALS patients 22 (22%) had a genetic mutation in one of the four major genes: 8 had mutation in *C9orf72*, 3 in TARDBP, 4 in *FUS* and 7 in *SOD1*. 7 MND had a concomitant FTD profile. No differences in sex and age between MND, MND mimics, and HCs were observed (p>0.05).

Demographic and clinical characteristics of the participants are summarized in **Table 3**.

**Table 3.** Participant demographic and clinical information.

|  | ALS<br>(100) | MND<br>mimics (73) | HCS<br>(38) | p-value |
| --- | --- | --- | --- | --- |
| Sex (M/F) | 60/40 | 44/29 | 20/18 | 0.93<sup>ξ</sup> |
| Age at T0 ± SD (y) | 64.5 ± 11.9 | 62.4 ± 13.9 | 64.2 ± 9.3 | 0.27<sup>ξ</sup> |
| Presence of genetic mutation | 21 | 13 | - | - |
| ALSFRSr_tot at diagnosis ± SD | 38.6 ± 5.8 | - | - | - |
| PR at diagnosis ± SD | 0.86 ± 0.74 | - | - | - |
| KINGS stage at diagnosis (0/1/2/3/4) | 0/27/25/44/4 | - | - | - |
| MiToS stage at diagnosis (0/1/2/3/4) | 78/14/8/0/0 | - | - | - |
| FVC ± SD<sup>α</sup> | 89.9 ± 20.8<sup>α</sup> | - | - | - |
| Diagnostic delay (months) | 11.2 ± 7.4 | - | - | - |

<sup>ξ</sup> $\chi^2$ test; <sup>α</sup> FVC values obtained around the data of blood sampling available in 74 ALS patients.
**Legend:** ALSFRSr\_tot: ALS Functional Rating Scale-revised total score; FVC: Forced Vital Capacity; SD: Standard Deviation; n/a: not assessed.

Plasma PRPH levels were significantly different (Kruskal-Wallis: chi-squared= 70.4, df=2, p=5.55×10^-16^; **Figure 1**) in MND patients (1.49 ± 0.63 ng/ml), MND mimics (0.79 ± 0.34 ng/ml) and HCs (0.94 ± 0.44 ng/ml). Post-hoc Bonferroni’s correction, revealed elevated PRPH in MND patients compared to MND mimics (p=5.33×10^-15^) and to HCs (p=2.55×10^-6^). These differences are still more pronounced if we exclude PLS and PMA and we consider only ALS (PRPH levels 1.53 ± 0.64 ng/ml; ALS vs MND mimics: chi-squared= 70.6, df=2, p=4.44×10^-16^). Looking at different types of MND (ALS, PLS, PMA), we did not observe statistical differences, although in PLS and PMA patients (1.26 ± 0.45 ng/ml) PRPH levels resulted slightly lower compared to ALS (1.53 ± 0.64 ng/ml). No differences between ALS/FTD and the other ALS patients (Mann-Whitney, p=0.483).

**Figure 1.**
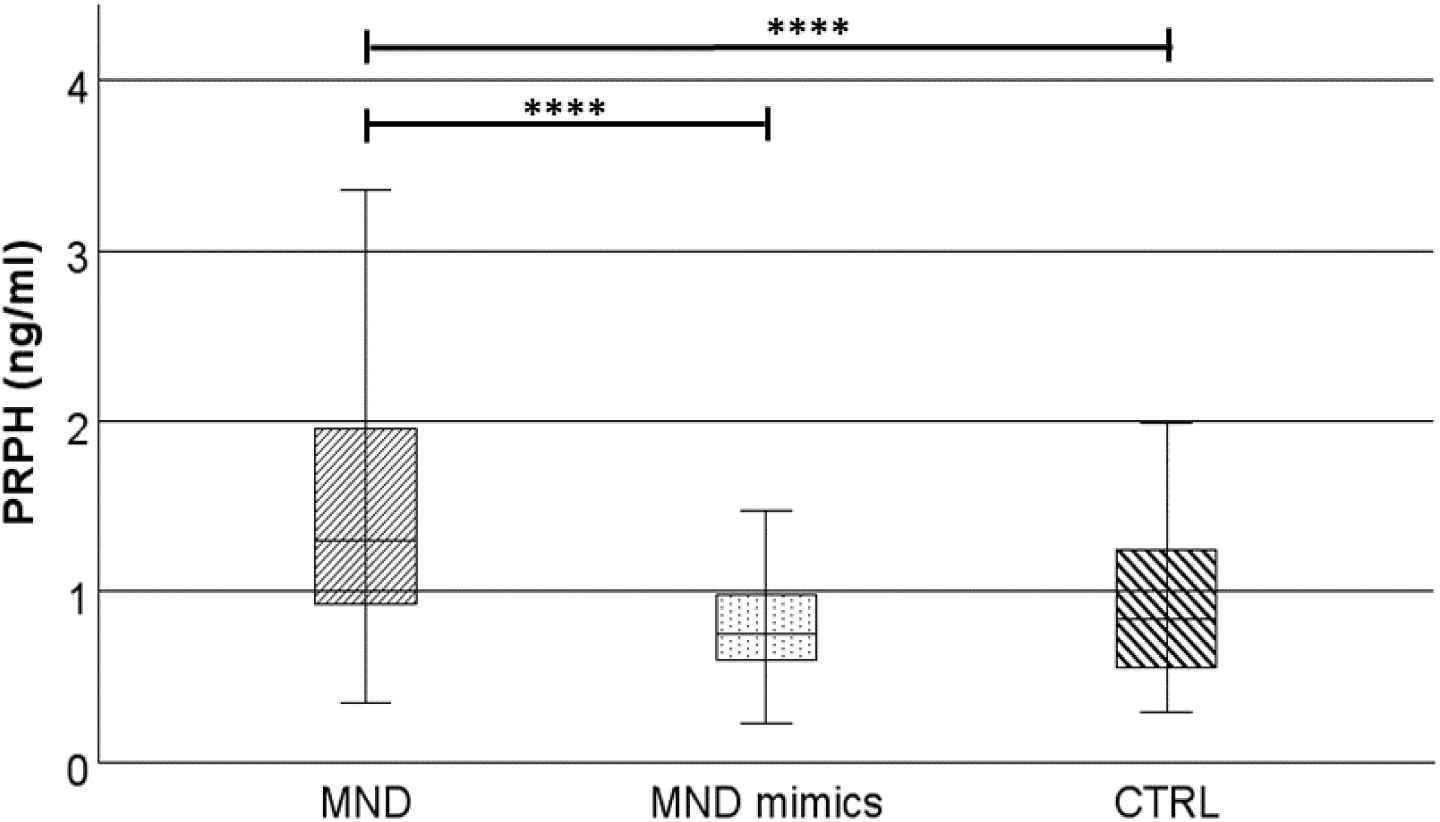
Levels of plasma PRPH in MND, MND-mimics, and HCs. <u>Legend</u>: PRPH= Peripherin; MND=motor neuron disease; HCs= healthy controls; ****= p≤0.0001. Box plots show median and interquartile range.

Based on these findings, we calculated the ROC curve for PRPH to differentiate ALS patients and MND mimics finding an AUC 0.846 (IC 0.791-0.902) and the best cut-off level of PRPH 1.06 ng/ml with a Youden index of 0.54, with a sensitivity of 0.770 and a specificity of 0.767 (see Supplementary data, Figure 5).

Moreover, we looked at the 15 PLS and at the 12 HSP included in our study, two overlap diseases with a similar onset and similar clinical characteristics during their course not easy to discriminate one each other without the genetic test. We observed that, PRPH levels were significantly higher (Mann Whitney, p=7.71×10) in PLS (1.35 ± 0.49 ng/ml) than in HSP (0.48 ± 0.25 ng/ml).

Within the ALS subgroup, patients harboring mutations in one of the four major known genes did not exhibit different levels of PRPH compared to those without genetic mutations (p>0.05).

No statistically significant differences in PRPH plasma levels were identified between ALS cases with bulbar and spinal onset (Mann-Whitney, p=0.91) nor among ALS with different phenotype nor between male and female patients (Mann-Whitney, p=0.98).

### Correlation of plasma PRPH levels with clinical, laboratory, and instrumental parameters in ALS patients

To reduce clinical heterogeneity, analyses focused on ALS patients. A modest positive correlation was noted between plasma PRPH levels and ALSFRSr scores at T0 (r_s_=0.364; p=0.0002; see **Figure 2A**), as well as ALSFRSr_noresp scores at T0 (r_s_=0.326; p=0.0009; see **Figure 2B**), ALSFRSr_4limbs at T0 (r_s_=0.261; p=0.009; see **Figure 2C**), ALSFRSr scores at T6 (r_s_=0.294; p=0.015; see **Figure 2D**), ALSFRSr_noresp scores at T6 (r_s_=0.257; p=0.035; see **Figure 2E**), and MRC at T6 (r_s_=0.266; p=0.034, see **Figure 2I**), while a negative correlation was observed between PRPH levels and PR at T0 (r_s_=- 0.225; p=0.02; see **Figure 2F**), and with LMNI at T0 (r_s_=-0.255; p=0.024; see **Figure 2G**). Moreover, PRPH levels at baseline seem to predict the trend of the disease in the next six months, calculated as the changes in progression rate (ΔPR%): a moderate inverse correlation was observed between PRPH at T0 and ΔPR% (r_s_=-0.343, p=0.006). All these results are confirmed after multiple regression analysis covarying for sex, age, and site of onset (**Supplementary data, Table 5**).

**Figure 2.**
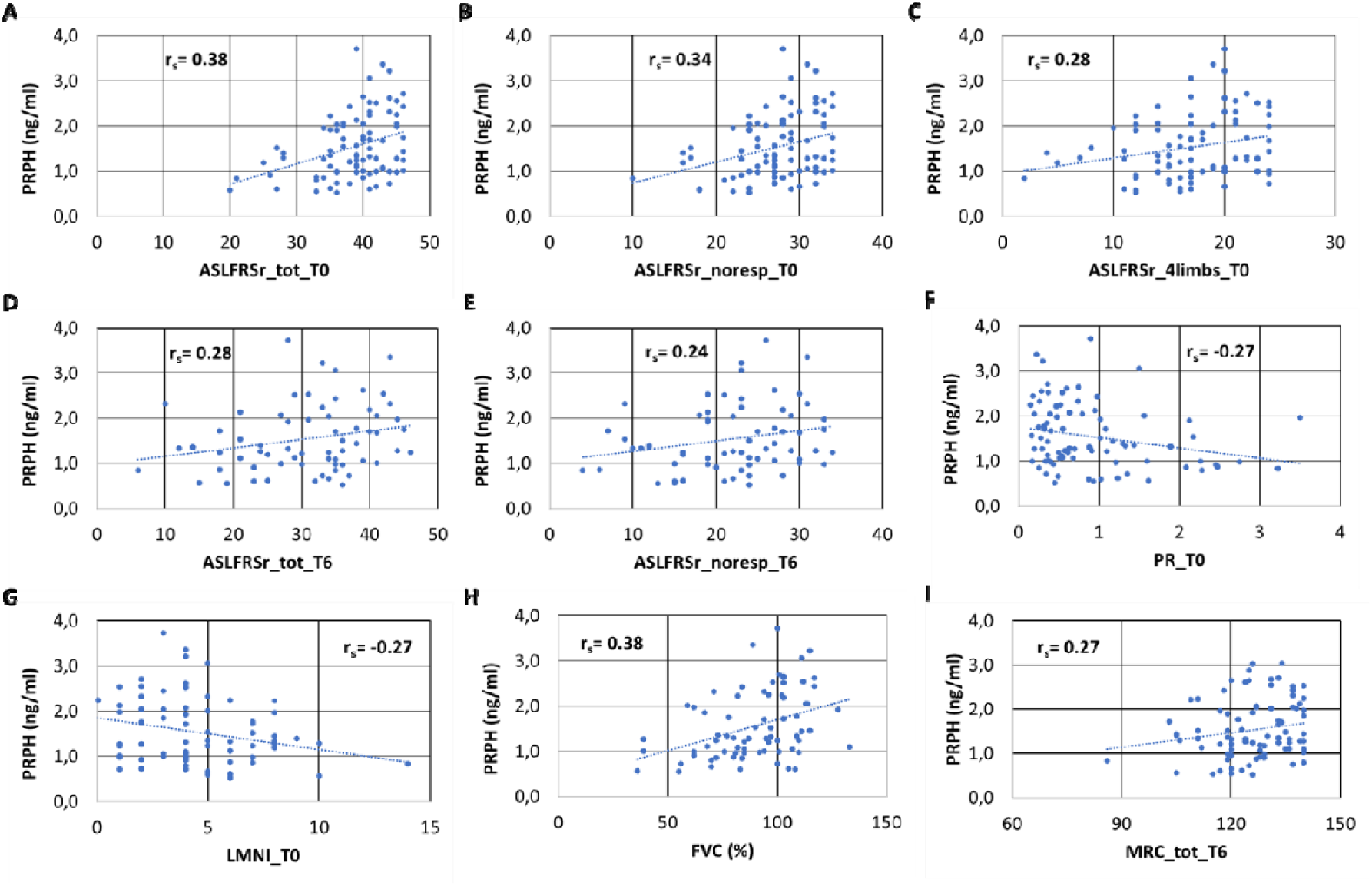
Correlation between clinical parameters and PRPH in ALS patients. <u>Legend</u>: PRPH= Peripherin; ALSFRSr_tot_T0: ALS Functional Rating Scale-revised total score at T0; ALSFRSr_noresp_T0: ALSFRSr without the respiratory item at T0; ALFRSr_4limb_T0: sum of item 4,5,6,7,8,9 of the ALSFRSr at T0; ALSFRSr_tot_T1: ALS Functional Rating Scale-revised total score at T1; ALSFRSr_noresp_T1: ALSFRSr without the respiratory item at T1; FVC: Forced Vital Capacity; LMNI: Lower Motor Neuron Index; MiToS: Milano-Torino staging system.

In support to the above reported evidences, we observed higher levels of plasmatic PRPH in ALS patients in early phases of disease, measured using the established MiToS and KINGS staging systems (ordinal logistic regression analysis: MiToS, Chi-square=7.897, p=0.005; KINGS, Chi-square=5.897, p=0.015). Covarying for age, sex, and site of onset the significance was confirmed and the model became stronger (MiToS, Chi-square=15.303, p_model=0.004, p_PRPH=0.01; KINGS, Chi-square=15.936, p_model=0.003, p_PRPH=0.026).

In a subset of 74 ALS patients in which it was available the value of forced vital capacity (FVC) collected around the data of blood sampling, a positive correlation was observed between FVC and PRPH (r_s_=0.382; p=0.0008; see **Figure 2H**). This result is confirmed after multiple regression analysis covarying for ALSFRSr at T0, sex, and age (**Supplementary data, Table 5**).

In contrast, no correlation was observed at either baseline or at six-month follow-up with PRPH levels and MRC score, change in MRC score (ΔMRC), UMNBS, duration of disease until blood sampling, or duration of disease between blood sampling and death or tracheostomy. Furthermore, we found no correlation between plasma PRPH levels and age at blood sampling.

### Survival analysis

Among the 100 ALS patients enrolled, 56 had either deceased or undergone tracheostomy by February 2024. Interim survival analyses were performed within this subgroup, acknowledging the limited sample size. Given the observed correlation between plasma PRPH levels, functional impairment, and disease progression, Kaplan-Meier survival curves were generated. PRPH levels were categorised using both the median value (≤1.35 ng/mL vs. >1.35 ng/mL) and tertiles (1st: 0.52–1.17 ng/mL; 2nd: 1.19–1.72 ng/mL; 3rd: 1.75–3.03 ng/mL), with tertile stratification demonstrating superior representation due to the log-normal distribution of PRPH data (Supplementary Data, Figure 6). Using the median-based stratification, a borderline statistically significant divergence in survival curves was observed (Log-Rank test, χ²=2.731, p=0.096; Breslow test, χ²=4.420, p=0.036; Tarone-Ware test, χ²=3.803, p=0.050, Figure 3A), which became more pronounced when stratified by site of onset (Log-Rank test, χ²=4.063, p=0.044; Breslow test, χ²=5.138, p=0.023; Tarone-Ware test, χ²=5.113, p=0.024). Tertile stratification revealed a significant increase in survival in patients with higher PRPH plasma levels (Log-Rank test, χ²=9.088, p=0.011; Breslow test, χ²=11.128, p=0.004; Tarone-Ware test, χ²=10.551, p=0.005; **Figure 3B**). This significance persisted when further stratified by site of onset (Log-Rank test, χ²=12.812, p=0.002; Breslow test, χ²=13.981, p<0.001; Tarone-Ware test, χ²=14.341, p<0.001).

**Figure 3.**
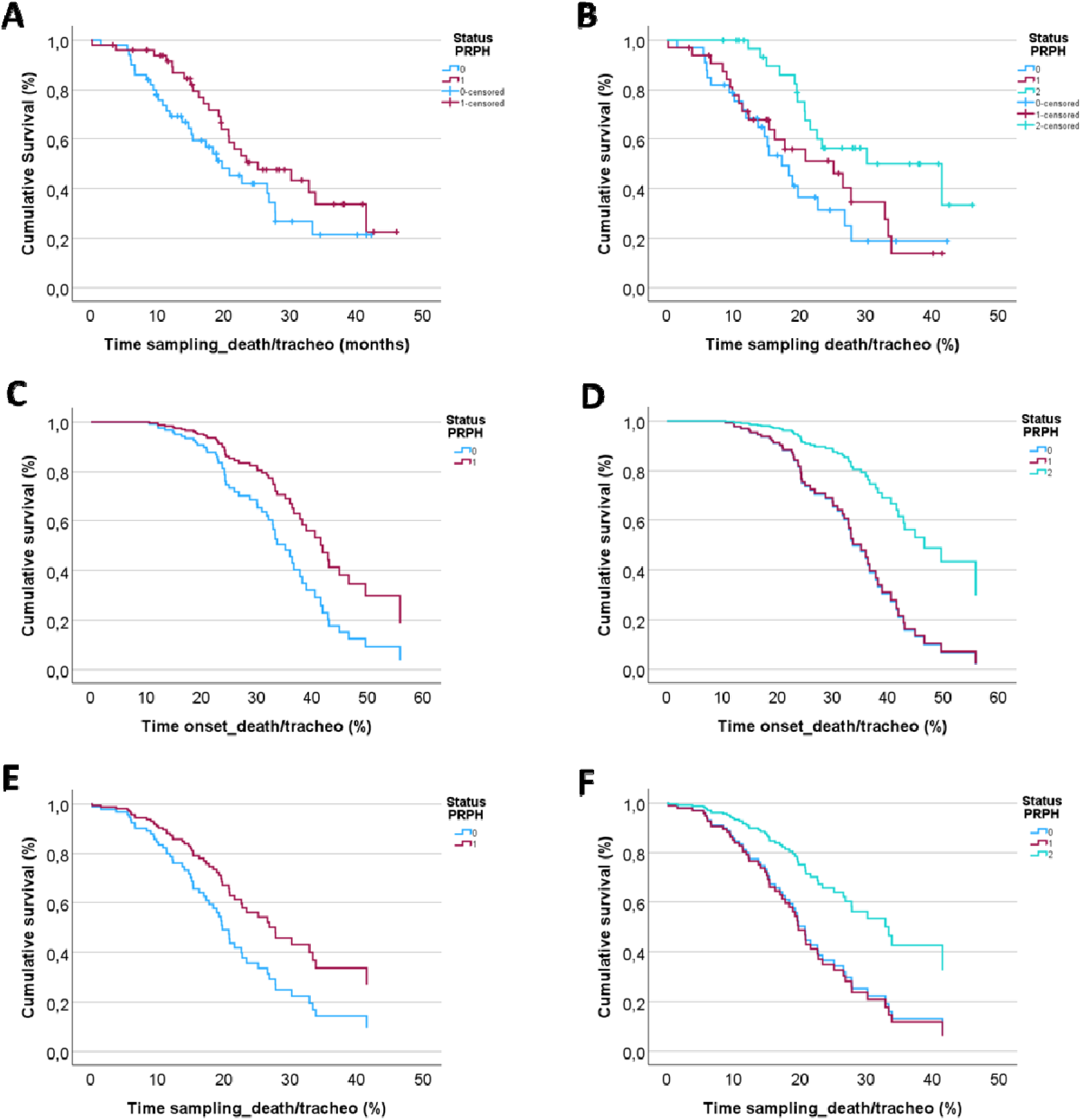
Survival analysis. (**A**) and (**B**) represent the Kaplan-Meier analysis, respectively categorising plasmatic PRPH levels using median and tertiles. (**C**) and (**D**) show survival curves covarying for plasmatic PRPH levels, sex, age at blood sampling, site of onset of the disease, and time from onset to death/tracheostomy, respectively categorising plasmatic PRPH levels using median and tertiles. (**E**) and (**F**) show survival curves covarying for plasmatic PRPH levels, sex, age at blood sampling, and site of onset of the disease, and time from blood sampling to death/tracheostomy, respectively categorising plasmatic PRPH levels using median and tertiles. <u>Legend</u>: in (A), (C), and (E) light-blue line (status PRPH 0) correspond to levels of PRPH below 1.35 ng/ml, while red line (status PRPH 1) to levels of PRPH above 1.35 ng/ml; in (B), (D), and (E) light-blue line (status PRPH 0) correspond to the 1° tertile (0.52-1.17 ng/ml), red line (status PRPH 1) to levels of PRPH 2° tertile (1.19-1.72 ng/ml), and light-green line (status PRPH 2) to levels of PRPH 3° tertile (1.75-3.03 ng/ml). Patients with higher levels of PRPH show a longer survival.

Univariate and multivariate Cox regression analyses were conducted for both continuous and categorical PRPH values at the time of blood sampling (**Supplementary Data, Table 6**). Elevated plasma PRPH levels were consistently associated with improved survival, particularly when using tertile stratification. These findings were confirmed for overall survival and survival post-blood sampling, using both continuous and categorical PRPH values. Graphical representations from multivariate Cox regression analyses both categorising for median and for tertiles are displayed below in **Figures 3C,D,E,F**.

### Longitudinal evaluation

No differences (Mann-Whitney p>0.1) are observed at baseline in terms of gender (M:F ratio 0.58 vs 0.54), age (64.5±11.9 vs 64.1±9.5), ALSFRSr (38.8±4.8 vs 38.3±5.7), PR (0.90±0.93 vs 0.96±0.98), MRC (126±11 vs 127±11), and FVC (89±20 vs 90±20) between the general group of 100 ALS patients and the subgroup of 41 subjects with ALS, for whom we measured PRPH levels approximately 6 months (mean: 6.68±1.67; median 6.37 [IQR 5.30–7.73]) after the initial collection. Major clinical parameters at T0 and at T6 of these 41 patients are shown in Supplementary data, Table 7. Analysing the longitudinal trend of plasma PRPH levels, no statistically significant variations are observed between T0 and T6 (Wilcoxon test, p=0.052), although the trend is decreasing (T0 1.63±0.73 ng/ml; T1 1.33±0.46 ng/ml; Figure 4).

**Figure 4.**
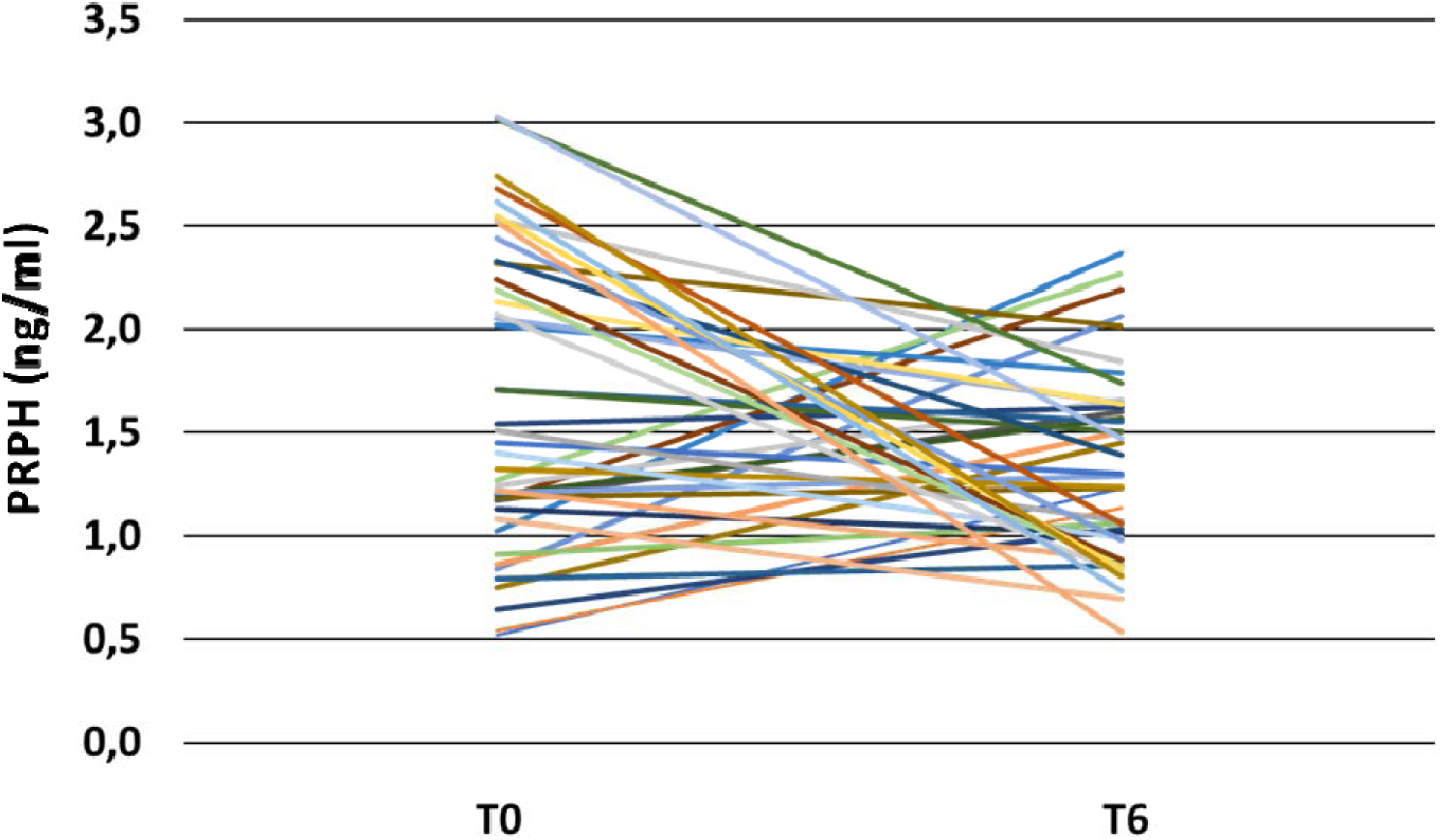
Longitudinal changing of plasma PRPH. <u>Legend</u>: PRPH= Peripherin; ΔPRPH%= percentage of change in PRPH levels between T0 and T6

## Discussion

In our study, we have examined the ability of plasma PRPH in distinguishing MND from MND- mimics and HCs. The most important finding is the satisfactory ability of PRPH to distinguish individuals affected by MND, and most of all by ALS, from those with MND-mimicking conditions, identifying, in our data, the optimal cut-off value as 1.06 ng/ml. These findings are in keeping with a previous study^11^, albeit their measurements were conducted in serum rather than plasma.

Drawing from these observations, the potential diagnostic utility of PRPH in patients in whom ALS is suspected is conceivable. Nevertheless, broader confirmation, including a concomitant comparison with other established neurofilaments, paralleled by adherence to standard laboratory techniques, remains essential, as reported in a previous study^12^.

Another promising application of PRPH lies in distinguishing PLS from HSP before genetic testing, which is time-consuming and costly. While these findings require larger multicentre validation, PRPH could improve early diagnostic efforts in pure upper motor neuron diseases.

Analysing the absolute levels of PRPH, we observed a substantial variability among the three available studies^11,12^, including the present one. These discrepancies may stem from variations in laboratory techniques, different kit employed, variances in sample processing methodologies, different atmospheric conditions, different dilutions, and distinctions in the type of material employed (plasma versus serum). Speculation may also extend to a potential blood matrix-effect resulting in the sequestration of PRPH in aggregates or its degradation by proteases or immunological binding, consequently reducing the free fraction, akin to what happen in plasma for NfL and pNfH^26^.

Focusing on ALS, plasma PRPH levels showed no associations with genetic factors, gender, age, ALS phenotype, disease site of onset, cognitive impairment, UMNS, and common blood examinations. In contrast to the findings reported in another study^11^, modest correlations were noted with clinical parameters. Specifically, higher PRPH levels correlated with less severe disease at the time of sampling (per staging systems), and better functional status (ALSFRSr, including sub-scores, and MRC). Furthermore, elevated plasma PRPH levels were associated with a lower disease progression rate prior to blood sampling and appear to predict a reduction in progression rate in the following months, independent of disease stage. Additionally, PRPH was inversely correlated with lower motor neuron damage both at the time of sampling and after six months, as assessed by the LMNI score aligning with its physiological concentration in the peripheral nervous system. An intriguing correlation between PRPH levels and FVC suggests that ALS patients with better-preserved respiratory function exhibit higher PRPH levels.

Survival analyses further supported PRPH’s prognostic value, showing that ALS patients with higher baseline PRPH levels had longer survival times, as evidenced by the graphs concerning both the time elapsed between onset and death/tracheostomy and that between sampling and death/tracheostomy. However, ongoing follow-up (nowadays, fortunately, 44 patients with ALS out of 100 are alive) and validation in larger multicentre studies are needed to solidify these findings.

All the above-mentioned data suggest that, unlike neurofilaments, which directly correlate with disease progression, number of regions affected, and motor neuron damage^27–29^, plasma PRPH levels may not indicate the extent of motor neuron impairment. Instead, PRPH levels could be an index of the intracellular repair attempts initiated by the motor neurons early in the disease. Elevated PRPH in early stages and its inverse relationship with disease progression contrast with typical neurodegenerative biomarkers. If this hypothesis holds true, PRPH levels could potentially increase as part of an upregulation process in response to neuronal injury, a phenomenon already observed in different cellular studies^30–32^. In particular, an elevation of PRPH blood levels has been described both in lesional models of the lower motor neuron and in genetic models of lower motor neuron neurodegeneration, and the level of PRPH correlate with the entity of lower motor neuron effort of biological repair.

Furthermore, the higher levels of PRPH in SBMA patients compared to ALS observed in another study^11^, support the idea of PRPH not merely as a marker of damage but as an indicator of mechanisms mitigating lower motor neuron damage. In fact, the less aggressive nature of SBMA, with slower progression rates, may explain this difference. Unfortunately, based on this study, we cannot determine the directionality of the causal effect: whether peripherin increases due to its involvement in the supposed repair attempt or its increase is merely an epiphenomenon of this process.

To clarify, when we talk about the attempt of repair and the mitigation of lower motor neuron damage, we do not mean a real clinical improvement of the disease, which is for definition relentlessly progressive, but merely we concern to a cellular compensatory mechanism. Future investigation of these mechanisms could open the field to a better understanding of rescue attempts of motor neurons in ALS and could let to identify possible targets of therapies.

However, such interpretations remain speculative: comprehensive and multicentre studies with extended follow-up periods, along with histological and neuropathological data and with confirmation in cellular and animal models, are necessary to validate these hypotheses.

The limitations of our study encompass the absence of additional fluid biomarkers concurrently measured (such as NfL and pNfH), the absence of ultrasensitive-techniques such as single-molecule array technology (SiMoA), the lack of validation evidence in preclinical models of ALS, the monocentric design of the study, and challenges in comparing these findings with the other two studies due to the utilization of plasma instead of serum in absence of a validated and standard protocol.

In conclusion, plasma levels of PRPH may serve as a useful biomarker in the differential diagnosis of ALS, in order to do an early diagnosis, and in prognosis definition. Further investigations, enrolling a larger patient cohort and employing more advanced PRPH quantification techniques, are undoubtedly necessary to validate our findings.

## Data Availability

All data produced in the present study are available upon reasonable request to the authors.

