## Supplementary data for "Peripherin: a novel early diagnostic and prognostic plasmatic biomarker in Amyotrophic Lateral Sclerosis"

**Table 4 – ALS mimics**

| **Diagnosis** | **Numerousness** |
| --- | --- |
| **Axonal polyneuropathies/mixed polyneuropathies** | 19 |
| **Hereditary Spastic Paraplegia** | 12 |
| **Cervical myelopathies/radiculopathy** | 5 |
| **Post-poliomyelitis** | 5 |
| **Benign fasciculations syndrome** | 5 |
| **FOSMN** | 4 |
| **Inflammatory myositis** | 3 |
| **Niemann-Pick type C** | 3 |
| **Primary Progressive Aphasia/Frontotemporal Dementia** | 3 |
| **Hirayama disease** | 2 |
| **Dyspnoea post-covid** | 2 |
| **Lumbar spinal stenosis** | 2 |
| **Miastenia gravis (bulbar onset)** | 2 |
| **Parsonage Turner** | 1 |
| **Progressive Supranuclear Palsy** | 1 |
| **Myofibrillar myopathies** | 1 |
| **Unilateral tongue atrophy** | 1 |
| **Functional disorder** | 1 |
| **Syringomyelia** | 1 |

**Table 5 – Multiple regression analysis between PRPH levels and clinical parameters**

| **Dependent variable ^δ^** | **Adjustments*** | **R adjusted** | **F** | **p-value** |
| --- | --- | --- | --- | --- |
| ***ALSFRSr_tot_T0*** | Age at blood sampling (cont), sex (M/F), site of onset (S/B) | 0.524 | 9.00 | **3.3 x10^-6** |
| ***ALSFRSr_noresp_T0*** |  | 0.510 | 8.34 | **8.2 x10^-6** |
| ***ALSFRSr_tot_T6 ^α^*** |  | 0.447 | 3.93 | **0.007** |
| ***ALSFRSr_noresp_T6 ^α^*** |  | 0.416 | 3.30 | **0.016** |
| ***PR_T0*** |  | -0.366 | 3.68 | **0.008** |
| ***ΔPR% ^α^*** |  | 0.487 | 4.43 | **0.003** |
| ***MRC_T6 ^α^*** |  | 0.576 | 7.09 | **0.0001** |
| ***LMNI_T0*** |  | 0.527 | 7.023 | **0.0001** |
| ***FVC ^ξ^*** | Age at blood sampling (cont), sex (M/F), site of onst (S/B), ALSFRSr_tot_T0 (cont) | 0.583 | 6.13 | **2.5 x10^-5** |

*Method used for inserting variables in the model: Stepwise Backwards

^δ^ analysis led on all the 100 ALS patients, except for α and ξ

^α^ analysis led on 68 ALS patients due to availability of clinical scal*e* data

^ξ^ analysis led on 74 ALS patients due to availability of spirometry data around the data of blood sampling

*Legend: ALSFRSr_tot: ALS Functional Rating Scale-revised total score; ALSFRSr_noresp: ALSFRSr without the respiratory items; PR: progression rate of disease; FVC: Forced Vital Capacity; T0: at the moment of sampling.*

**Table 6 – Cox proportional hazard models**

| **Univariate Analysis—Overall Survival** | | | | |
| --- | --- | --- | --- | --- |
|  | | | **HR (95% CI)** | **p** |
| **Age at blood sampling (continuous)** | | | 1.033 (1.006-1.061) | **0.016** |
| **Sex** | | | 0.912 (0.529-1.572) | 0.741 |
| **Site of onset (S/B)** | | | 2.623 (1.516-4.539) | **0.0005** |
| **Time onset to blood sampling** | | | 0.929 (0.896-0.963) | **6.91 x10^-5** |
| **PRPH (categorial-upper/lower median)** | | | 0.694 (0.407-1.182) | 0.179 |
| **PRPH (categorial)** | | **1° tertile**  **(<1.17 ng/ml)** | 1 |  |
|  |  | **2° tertile**  **(1.19-1.72 ng/ml)** | 0.780 (0.422-1.444) | 0.429 |
|  |  | **3° tertile**  **(1.75-3.03 ng/ml)** | 0.443 (0.224-0.877) | **0.019** |
| **PRPH (continuous)** | | | 0.637 (0.418-0.972) | **0.037** |
| **Multivariate analysis*—Overall survival**  *Adjusting for PRPH, age (cont), sex (M/F), site of onset (S/B),*  *time onset_blood sampling (cont)* | | | | |
|  | **Statistic of the model - PRPH (categorial - median)** | | | **2.06 x10^-6** |
|  | **PRPH (median)** | | 0.509 (0.288-0.899) | **0.020** |
| **Statistic of the model - PRPH (categorial - tertile)** | | | | **4.51 x10^-7** |
| **PRPH (categorial)** | | **1° tertile**  **(<1.17 ng/ml)** | 1 |  |
|  |  | **2° tertile**  **(1.19-1.72 ng/ml)** | 0.982 (0.505-1.909) | 0.958 |
|  |  | **3° tertile**  **(1.75-3.03 ng/ml)** | 0.311 (0.150-0.644) | **0.002** |
| **Statistic of the model - PRPH (continuous)** | | | | **1.77 x10^-7** |
| **PRPH (continuous)** | | | 0.554 (0.359-0.856) | **0.008** |
| **Univariate Analysis—Survival after sampling** | | | | |
|  | | | **HR (95% CI)** | **p** |
| **Age at blood sampling** | | | 1.032 (1.005-1.059) | **0.020** |
| **Sex** | | | 0.962 (0.561-1.652) | 0.889 |
| **Site of onset (S/B)** | | | 2.589 (1.479-4.532) | **0.0009** |
| **PRPH (categorial-upper/lower median)** | | | 0.640 (0.376-1.091) | 0.101 |
| **PRPH (categorial)** | | **1° tertile**  **(<1.17 ng/ml)** | 1 |  |
|  |  | **2° tertile**  **(1.19-1.72 ng/ml)** | 0.788 (0.426-1.457) | 0.447 |
|  |  | **3° tertile**  **(1.75-3.03 ng/ml)** | 0.369 (0.186-0.730) | **0.004** |
| **PRPH (continuous)** | | | 0.574 (0.379-0.870) | **0.009** |
| **Multivariate analysis*—Survival after sampling**  *Adjusting for PRPH, age (cont), sex (M/F), and site of onset (S/B)* | | | | |
| **Statistic of the model - PRPH (categorial - median)** | | | | **0.0003** |
| **PRPH (median)** | | | 0.563 (0.328-0.967) | **0.037** |
| **Statistic of the model - PRPH (categorial - tertile)** | | | | **2.71 x10^-5** |
| **PRPH (categorial)** | | **1° tertile**  **(<1.17 ng/ml)** | 1 |  |
|  |  | **2° tertile**  **(1.19-1.72 ng/ml)** | 1.166 (0.608-2.236) | 0.644 |
|  |  | **3° tertile**  **(1.75-3.03 ng/ml)** | 0.369 (0.186-0.733) | **0.004** |
| **Statistic of the model - PRPH (continuous)** | | | | **5.00 x10^-5** |
| **PRPH (continuous)** | | | 0.568 (0.384-0.840) | **0.005** |

**Method used for inserting variables in the model: Stepwise Backwards Regression*

**^α^***For MiToS: ‘low’ correspond to score 0 or 1, ‘high’ correspond to score ≥2; For KINGS: ‘low’ correspond to score ≤2, ‘high’ correspond to score ≥ 3*

**Table 7 – Clinical parameters and scales in ALS patients who underwent longitudinal sampling**

|  | **ALS (41)** | |  |
| --- | --- | --- | --- |
|  | **T0** | **T1** | **p-value**  **(*Wilcoxon test*)** |
| **ALSFRSr_tot ± SD** | 38.6 ± 5.8 | 29.9 ± 9.6 | **7.76 x 10^-7^** |
| **ALSFRSr_noresp ± SD** | 27.4 ± 5.0 | 21.3 ± 7.4 | **1.14 x 10^-5^** |
| **PR ± SD** | 0.88 ± 0.86 | 1.28 ± 0.97 | **0.003** |
| **PR_noresp ± SD** | 0.80 ± 0.74 | 0.91 ± 0.74 | 0.339 |
| **MRC_tot ± SD** | 126 ± 11 | 109 ± 26 | **1.72 x 10^-6^** |
| **FVC ± SD** | 90 ± 20 | n/a | n/a |
| **UMNBS ± SD** | 9.3 ± 4.7 | 9.2 ± 4.7 | **0.011** |
| **LMNI ± SD** | 4.9 ± 2.6 | 7.5 ± 3.3 | **4.84 x 10^-6^** |

*Legend: ALSFRSr_tot: ALS Functional Rating Scale-revised total score; ALSFRSr_noresp: ALSFRSr without the respiratory items; PR: progression rate of disease; PR_noresp: PR without considering the respiratory items; MRC_tot: total Medical Research Council scale; ΔMRC: monthly loss of point at MRC scale; UMNBS: Upper Motor Neuron Burden Score; LMNI: Lower Motor Neuron Index; FVC: Forced Vital Capacity; T0: at the moment of sampling; T6: after 6 months from sampling; n/a: not assessed.*

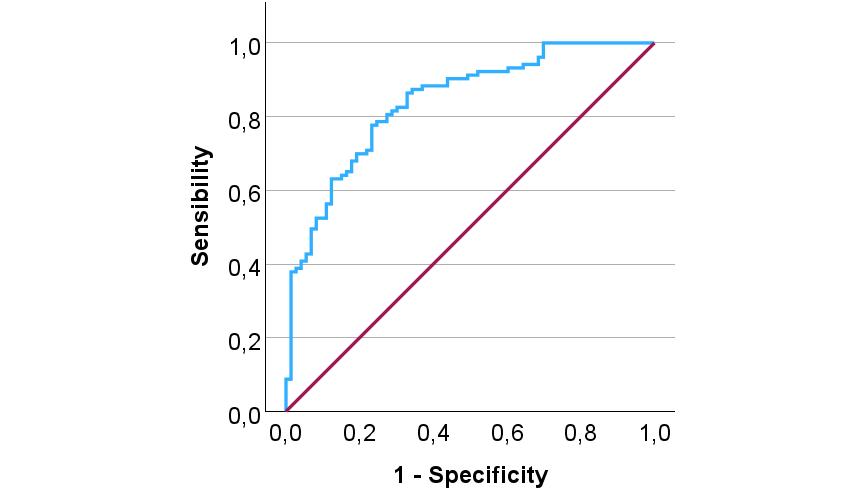

**AUC: 0.846**

**Figure 5 - ROC Curve comparing plasma levels of PRPH in ALS and MND-mimics.**

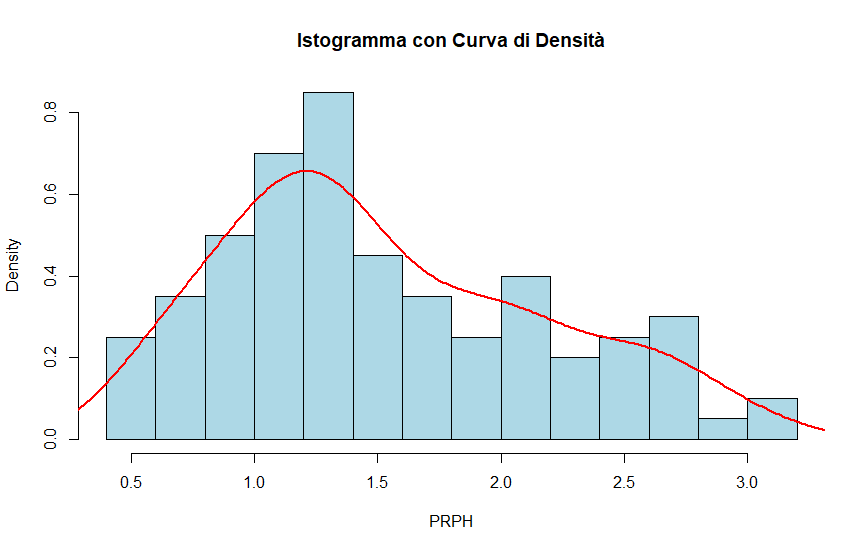

**Figure 6 - The log-normal distribution of PRPH data in ALS.**

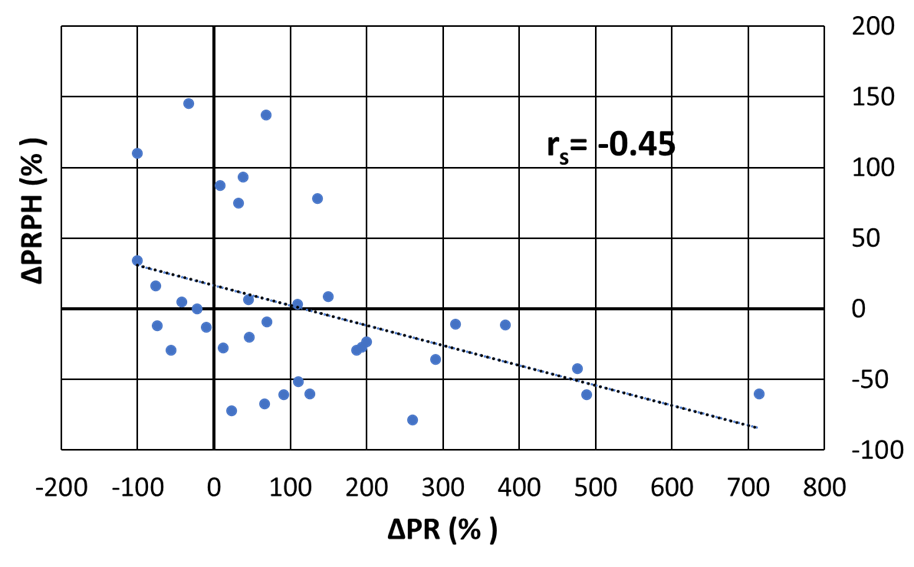

**Figure 7 – Longitudinal changing of plasma PRPH and correlation with progression of disease**

To better evaluate the possible role of plasmatic PRPH as a dynamic index of the attempt of the lower motor neuron to mitigate the neurodegenerative damage, we calculated the percentage change in PRPH levels between T0 and T6 (*ΔPRPH%*) and the percentage change in PR values between T0 and T6 (*ΔPR%*). A moderate inverse correlation was observed between ΔPRPH% and ΔPR% (r_s_= -0.452, p= 0.008; **Figure 4B**). Since we previously reported that PRPH plasmatic levels are influenced by the stage of the disease in ALS, we performed a backward stepwise multiple linear regression analysis, covarying for MiToS score at T0, age at T0, and sex. Despite the small sample size, the significance of this correlation was maintained (r_adjusted_= -0.530; F= 5.852, p= 0.007).

*Legend: PRPH= Peripherin; ΔPRPH%= percentage of change in PRPH levels between T0 and T6; ΔPR%= percentage of change in PR values between T0 and T6; T0: at the moment of sampling; T6: after 6 months from sampling.*
